# Multi-omics proposes a complement-adhesion-stromal-vascular (CASVR) framework in chronic rhinosinusitis with nasal polyps

**DOI:** 10.64898/2026.10.13.26365478

**Authors:** Zi-Ang Ma, Xiang Gao, JinLiang Yu, Yu Song, Junxiu Liu

**Author notes:** Correspondence (Y.S.); (J.L.).

## Abstract

Type 2 inflammation is central to chronic rhinosinusitis with nasal polyps (CRSwNP), but it does not fully explain persistent stromal and vascular remodeling. We asked whether complement, adhesion, stromal, and vascular signals converge across tissue, cellular, and spatial scales. We integrated four bulk-transcriptomic cohorts with a single-cell atlas and GeoMx spatial transcriptomics. Network analysis identified eight contrast-associated hub genes and an ABCC9-ACTA2-GPM6A score that separated study-defined groups across cohorts. Single-cell mapping localized these genes mainly to pericytes, fibroblasts, and endothelial cells, while CellChat linked structural and immune compartments through extracellular-matrix and inflammatory signals. In spatial data, C3-C3AR1 showed closer-than-random proximity after multiple-testing correction, whereas ICAM1-ITGAL/ITGB2 did not. Anatomical-site sensitivity analyses retained all eight hub-gene directions. These findings support CASVR as an association framework linking complement-related signals to structural-vascular compartments and identify C3-C3AR1 for independent mechanistic testing.

## Introduction

Chronic rhinosinusitis with nasal polyps (CRSwNP) affects approximately 2–4% of adults and substantially impairs quality of life [1]. Intranasal corticosteroids, endoscopic sinus surgery, and biologics can improve disease control, but treatment selection and durable responses remain difficult [1]. Disease recurrence and incomplete treatment responses suggest that inflammation is coupled to persistent changes in tissue structure [1,2].

Type 2 (T2) inflammation, characterized by IL-4, IL-5, IL-13, IgE production, and eosinophilia, provides the dominant mechanistic and therapeutic framework for CRSwNP [2,3]. T2-high and T2-low classifications, however, do not capture geographical heterogeneity, unequal responses to biologics, or progressive tissue remodeling [2–6]. Vascular dysfunction, pericyte– fibroblast alterations, and immune-cell retention are particularly difficult to explain using a cytokine-centered account alone. Their relationship to complement activation and leukocyte adhesion in CRSwNP tissue remains uncertain.

Single-cell studies have identified basal-cell–immune interactions, epithelial-to-fibroblast transitions, IL-1β-driven neutrophil recruitment, and broader epithelial–immune remodeling in CRSwNP [7,8]. Spatial transcriptomic platforms can place these cell-state and communication signals within tissue architecture [7–9]. Separate studies have reported transcriptomic classifiers, single-cell alterations, and proteomic recurrence markers [10–12], but these evidence streams have largely been examined independently. Whether complement- and adhesion-related programs are associated with stromal-vascular remodeling across bulk, single-cell, and spatial CRSwNP datasets remains unclear. The cell compartments carrying these signals, and whether prioritized ligand–receptor axes show nonrandom spatial proximity, are also unresolved.

Here, we asked whether complement, adhesion, and stromal-vascular signals form a coherent cross-scale association in CRSwNP. We first integrated multi-cohort bulk transcriptomes to identify group-associated pathways and networks and to derive a parsimonious transcriptomic score. We then used single-cell data to localize the selected genes and infer structural–immune communication. Finally, image-aware GeoMx digital spatial profiling data were used to map candidate genes and test area-of-interest proximity for selected ligand–receptor axes. We use complement-adhesion-stromal-vascular remodeling (CASVR) as a descriptive framework for these associations (Figure 1). Because C3AR1 and ABCC9 connected the complement and structural-vascular arms of CASVR, we also evaluated whether available chemical and structural evidence could support their pharmacological follow-up.

**Figure 1.**
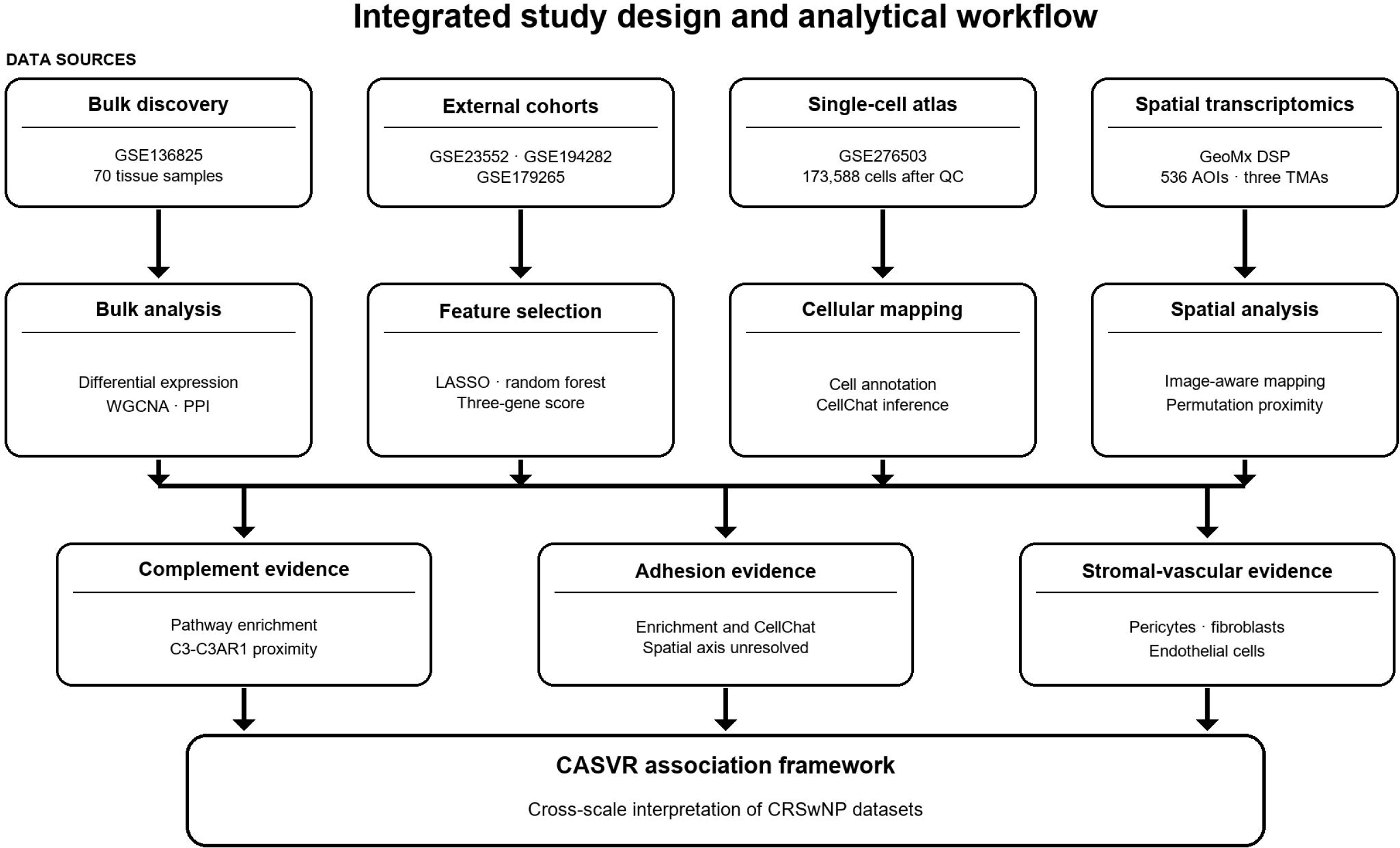
Integrated study design and analytical workflow. Bulk discovery data were used for differential expression, network analysis, and feature selection, with three external cohorts used for cross-cohort assessment. The GSE276503 single-cell atlas localized selected genes and supported CellChat inference. GeoMx digital spatial profiling mapped target genes and tested ligand–receptor proximity. The integrated evidence was organized into the CASVR association framework. Abbreviations: CASVR, complement-adhesion-stromal-vascular remodeling; DEG, differentially expressed gene; WGCNA, weighted gene co-expression network analysis; PPI, protein-protein interaction; LASSO, least absolute shrinkage and selection operator; AOI, area of interest; TMA, tissue microarray.

## Results

### Bulk transcriptomes reveal complement and vascular-wall signals

The GSE136825 contrast identified 562 differentially expressed genes, with 370 higher and 192 lower in CRSwNP nasal-polyp tissue than in control inferior-turbinate tissue (Figure 2a,b). Gene-set enrichment [13] highlighted immunoregulatory interactions, complement, and vascular-wall programs (Figure 2c-f). Complement and vascular-wall pathways remained among the strongest enriched programs after multiple-testing correction. Because the primary tissue groups differed by anatomical site, these findings describe the tissue-group contrast rather than a disease-specific effect.

**Figure 2.**
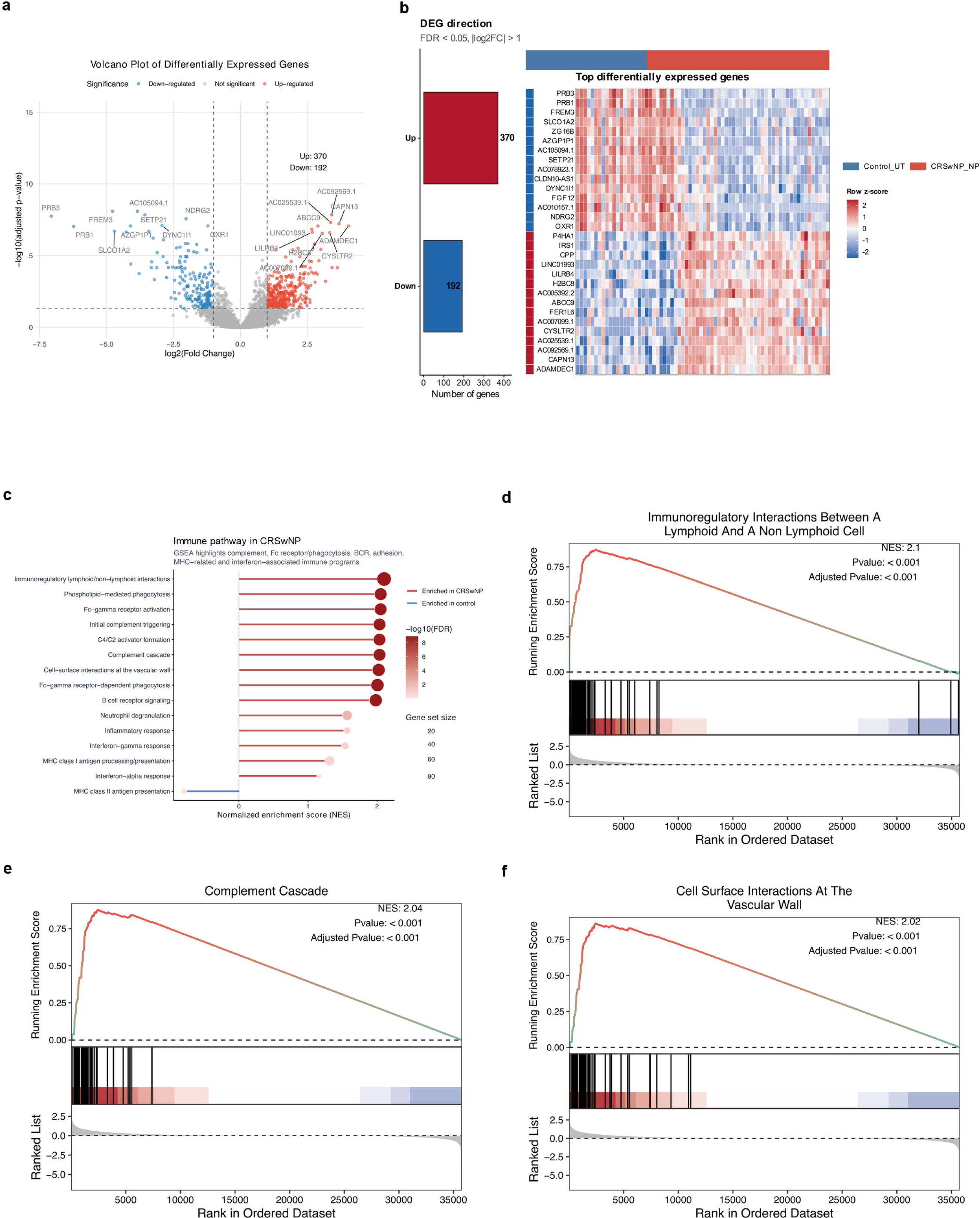
Bulk transcriptomic and pathway differences in the CRSwNP_NP-versus-Control_UT contrast. (a) Volcano plot of 370 higher and 192 lower genes at FDR < 0.05 and |log2FC| > 1 in GSE136825. (b) DEG counts and row-z-score heatmap of representative genes. (c) GSEA overview of enriched immune and structural pathways. Enrichment plots show (d) immunoregulatory interactions between lymphoid and non-lymphoid cells, (e) the complement cascade, and (f) cell-surface interactions at the vascular wall. Abbreviations: CRSwNP_NP, CRSwNP nasal-polyp tissue; Control_UT, control inferior-turbinate tissue; DEG, differentially expressed gene; FDR, false discovery rate; GSEA, gene set enrichment analysis; NES, normalized enrichment score.

### Network convergence yields a structural-vascular gene set and three-gene score

Five group-associated WGCNA modules contributed 396 candidate genes. Their intersection with the 562 differentially expressed genes yielded 208 genes for protein-interaction analysis (Figure 3a,b). Five cytoHubba rankings converged on NTRK3, GPM6A, SLITRK5, SPARCL1, ACTA2, RERG, ABCC9, and PDPN (Figure 3c-e). ABCC9, RERG, and PDPN were higher, whereas SLITRK5, GPM6A, ACTA2, NTRK3, and SPARCL1 were lower in the primary contrast.

**Figure 3.**
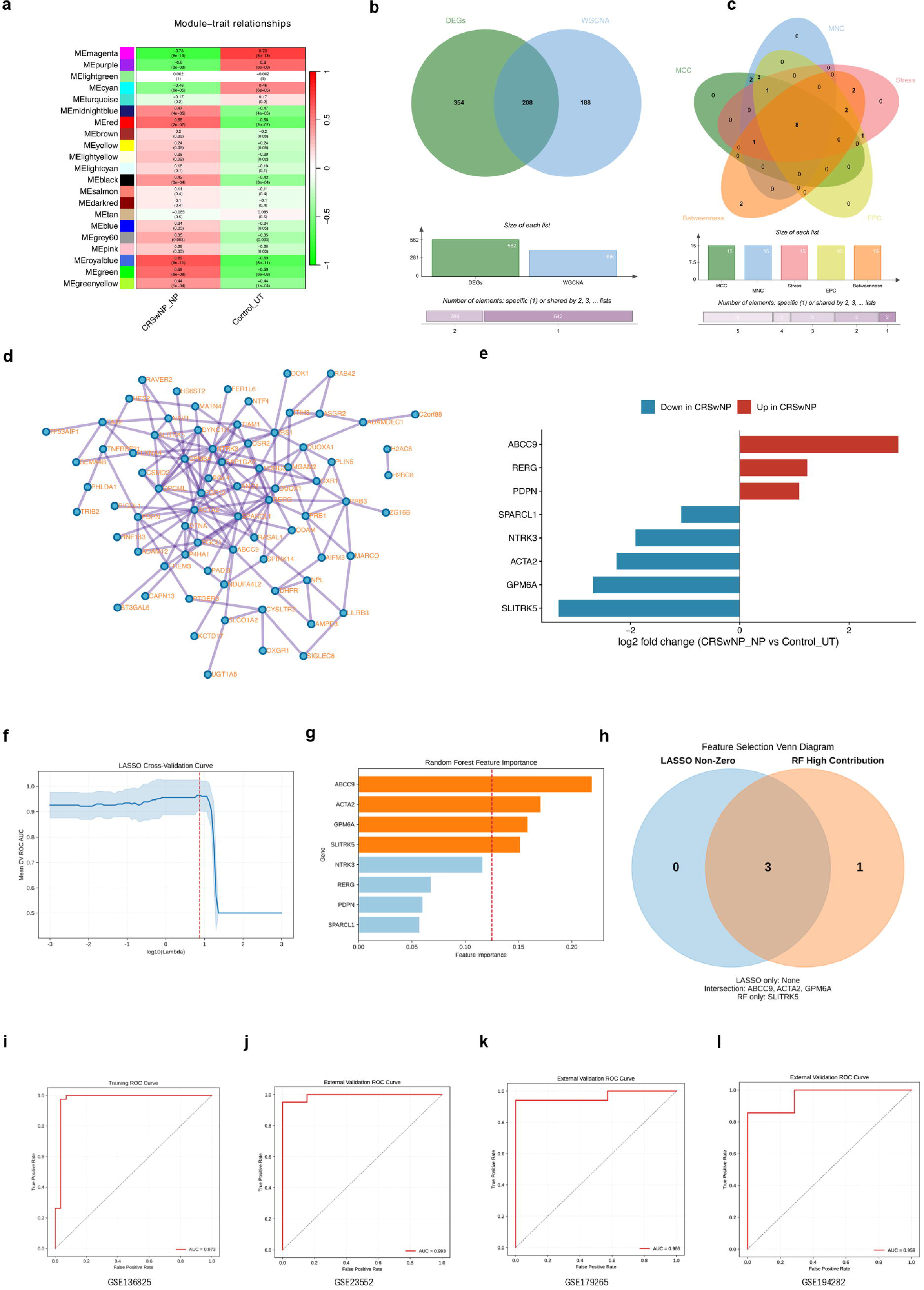
Network convergence and development of the ABCC9-ACTA2-GPM6A score. (a) WGCNA module-phenotype correlations identified group-associated modules. (b) Intersection of 562 DEGs with 396 WGCNA candidates yielded 208 genes. (c) The intersection of five cytoHubba rankings (MCC, MNC, EPC, Stress, and Betweenness) identified NTRK3, GPM6A, SLITRK5, SPARCL1, ACTA2, RERG, ABCC9, and PDPN. (d) STRING network of the 208 genes. (e) Hub-gene log2 fold changes for CRSwNP_NP versus Control_UT: ABCC9, RERG, and PDPN were higher, whereas SLITRK5, GPM6A, ACTA2, NTRK3, and SPARCL1 were lower. (f) LASSO retained ABCC9, ACTA2, and GPM6A. (g) Random-forest importance ranked ABCC9, ACTA2, GPM6A, and SLITRK5 above the mean. (h) The intersection of LASSO and random forest yielded the three-gene score. (i) Apparent ROC performance in GSE136825 (AUC 0.973). (j–l) Sample-level cross-cohort AUCs were 0.993 (GSE23552), 0.966 (GSE179265), and 0.959 (GSE194282). Abbreviations: WGCNA, weighted gene co-expression network analysis; DEG, differentially expressed gene; PPI, protein-protein interaction; LASSO, least absolute shrinkage and selection operator; ROC, receiver operating characteristic; AUC, area under the receiver operating characteristic curve.

LASSO retained ABCC9, ACTA2, and GPM6A; random forest also retained these genes together with SLITRK5 (Figure 3f-h). A score refitted from the three-gene intersection separated the study-defined groups in GSE136825 (apparent AUC, 0.973) and three external cohorts, whose sample-level AUCs ranged from 0.959 to 0.993 (Figure 3i-l and Supplementary Table S6).

Same-site and public-title-matched paired analyses preserved all eight hub-gene directions (Supplementary Fig. S1 and Supplementary Tables S1-S3). Sensitivity results for the distinct 35-gene CASVR module score are reported in the Supplementary Information.

### Single-cell data localize the gene set to structural and vascular compartments

After quality control and integration, the GSE276503 atlas contained 173,588 cells spanning epithelial, immune, stromal, and vascular populations (Figure 4a,b). ABCC9 localized mainly to pericytes and fibroblasts, ACTA2 to pericytes and contractile stromal cells, and GPM6A to endothelial cells (Figure 4c-e). The other hub genes showed related endothelial, stromal, and pericyte distributions.

**Figure 4.**
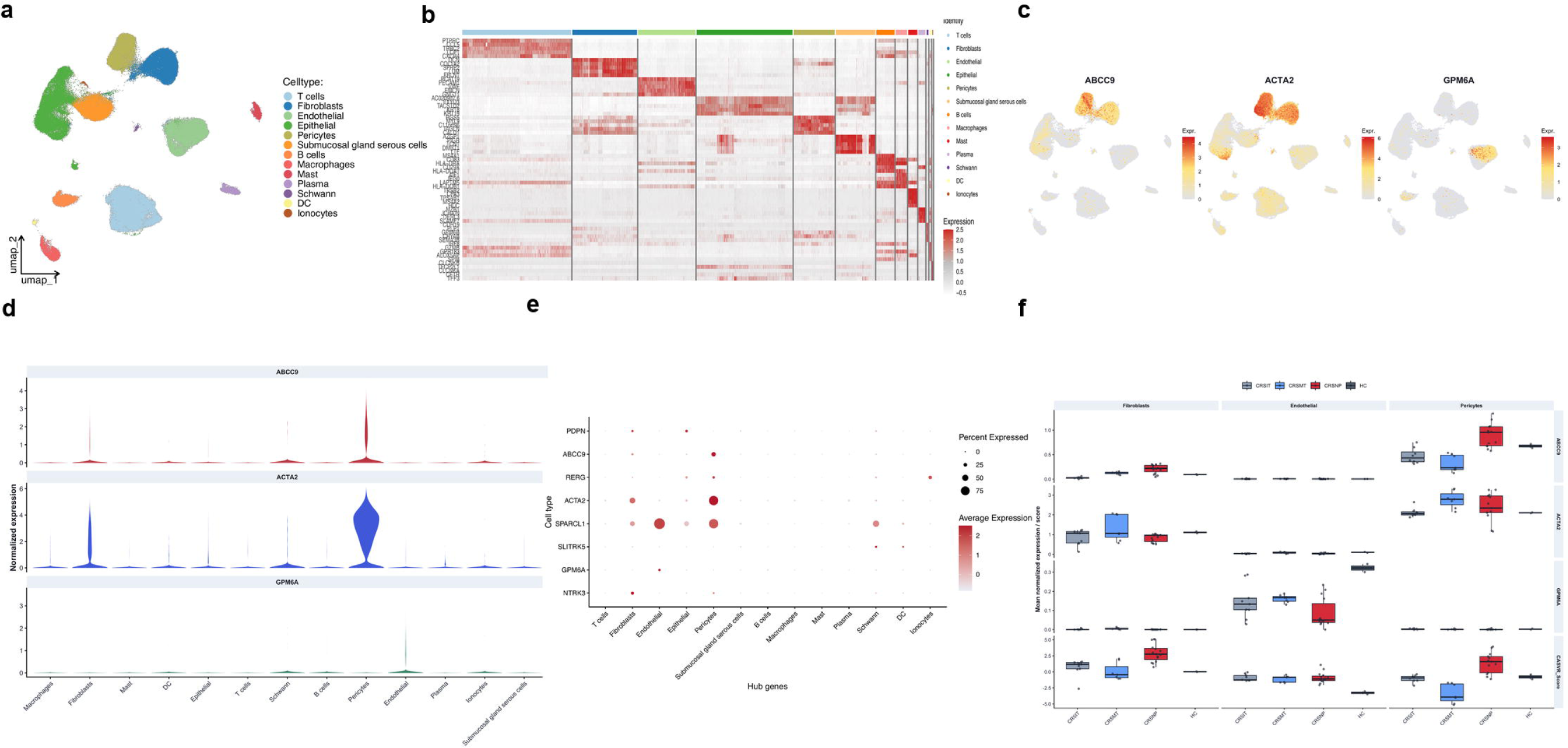
Single-cell localization of the network genes. (a) UMAP of 173,588 quality-controlled cells annotated into 13 cell types. (b) Marker-gene heatmap supporting the annotations. (c,d) Feature and violin plots localizing ABCC9 and ACTA2 mainly to pericytes and fibroblasts, and GPM6A mainly to endothelial cells. (e) Dot plot of the eight hub genes across cell types. (f) ABCC9, ACTA2, GPM6A, and CASVR scores across sampling groups within fibroblasts, endothelial cells, and pericytes. Abbreviations: UMAP, uniform manifold approximation and projection; CASVR, complement-adhesion-stromal-vascular remodeling; HC, healthy control.

### CellChat identifies candidate routes of structural-immune communication

CellChat inferred 2,771 interactions among the annotated populations (Figure 5a). Collagen, laminin, FN1, MIF, APP, and antigen-presentation outgoing and incoming signaling patterns differed between structural and immune populations (Figure 5b-e). The displayed sender-receiver pairs included APP-CD74, COL1A2-CD44, MIF-CD74, and MIF-CXCR4.

**Figure 5.**
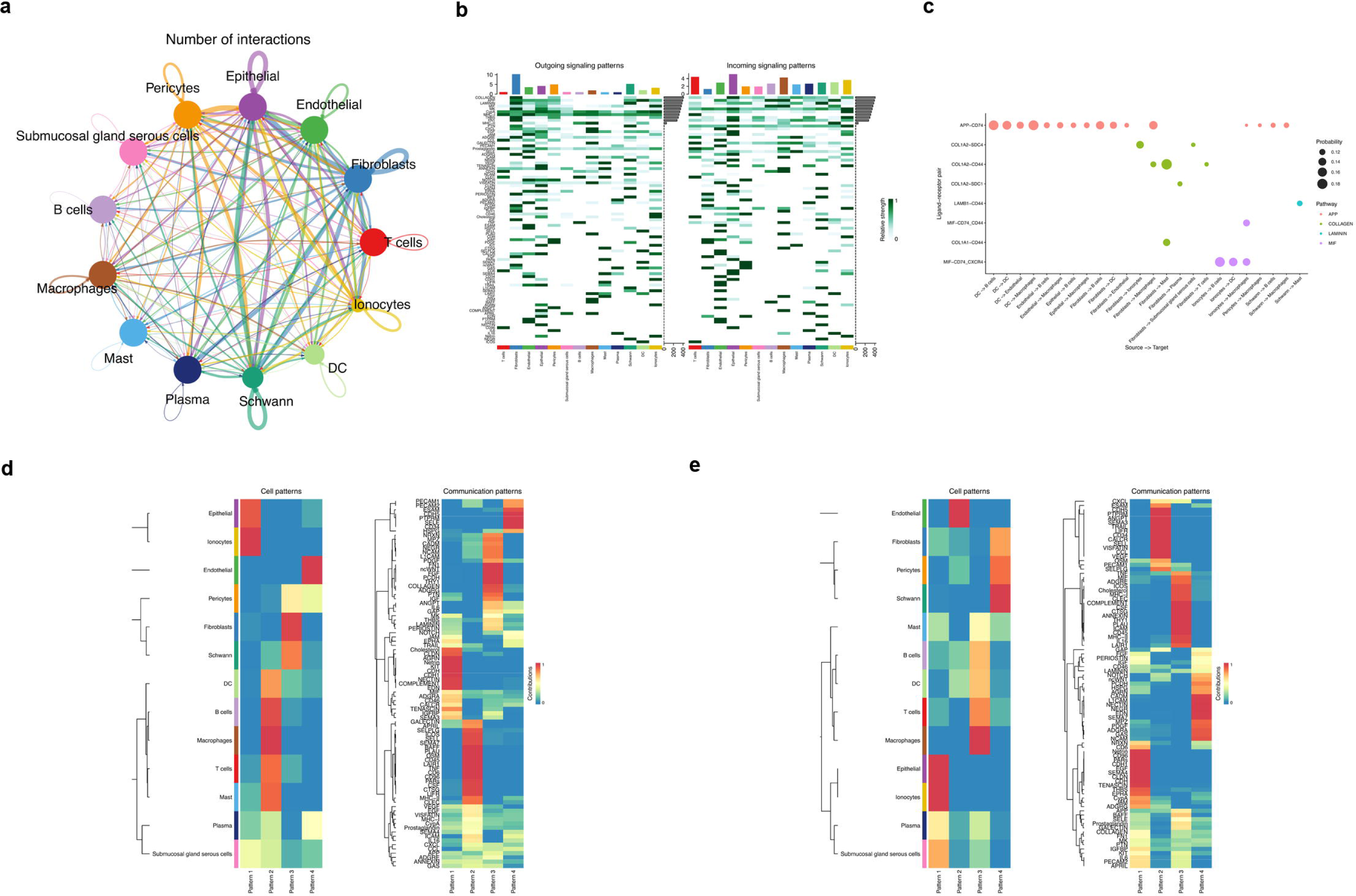
CellChat-inferred communication in the integrated single-cell dataset. (a) Total inferred interactions among annotated cell types. (b) Outgoing and incoming patterns for extracellular-matrix, inflammatory, and antigen-presentation pathways. (c) Selected sender-receiver pairs, including APP-CD74, COL1A2-CD44, MIF-CD74, and MIF-CXCR4. (d,e) Cell populations assigned to outgoing and incoming communication patterns. These panels report computational predictions for experimental testing. Abbreviations: ECM, extracellular matrix; MIF, macrophage migration inhibitory factor; APP, amyloid beta precursor protein; MHC, major histocompatibility complex.

### Spatial mapping prioritizes C3-C3AR1 and ICAM1-ITGAL/ITGB2 proximity

Spatial mapping used 536 retained areas of interest across three tissue microarrays. Hub-gene expression varied across tissue microarrays (Figure 6a), as did the predominant epithelial, immune, and macrophage-enriched segment classes (Figure 6b).

**Figure 6.**
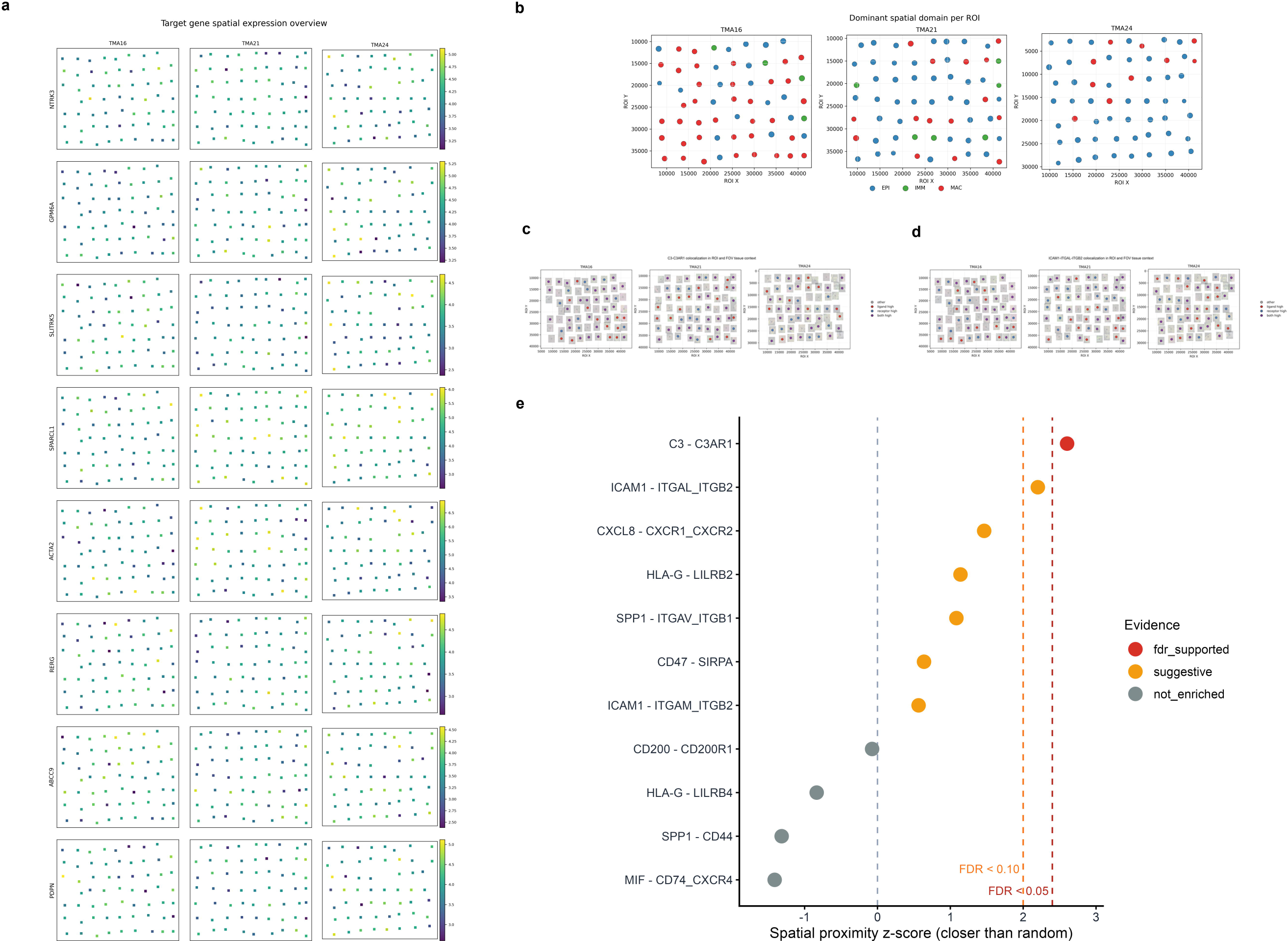
Image-aware spatial mapping and upper-quartile ligand–receptor proximity analysis. (a) Spatial expression overview of NTRK3, GPM6A, SLITRK5, SPARCL1, ACTA2, RERG, ABCC9, and PDPN. (b) Predominant GeoMx segment classes across ROIs in TMA16, TMA21, and TMA24. (c) Distribution of C3-high, C3AR1-high, and both-high AOIs at the upper-quartile cutoff. (d) Corresponding map for ICAM1 and the ITGAL/ITGB2 receptor score. The maps in c and d provide tissue context but do not establish statistical colocalization. (e) Ranking of 11 candidate ligand–receptor axes by closer-than-random proximity z-score; C3-C3AR1 was the only axis that met FDR < 0.05. Abbreviations: TMA, tissue microarray; ROI, region of interest; AOI, area of interest; EPI, epithelial segment; IMM, immune segment; MAC, macrophage-enriched segment; LR, ligand–receptor; FDR, false discovery rate.

At the upper-quartile expression cutoff, C3-C3AR1 had the highest closer-than-random proximity score among 11 candidate axes and passed multiple-testing correction (Figure 6c,e). ICAM1-ITGAL/ITGB2 ranked second but did not pass this threshold (Figure 6d,e). Results for all axes are reported in Supplementary Table S4. These cross-scale relationships are summarized in the CASVR framework (Figure 7).

**Figure 7.**
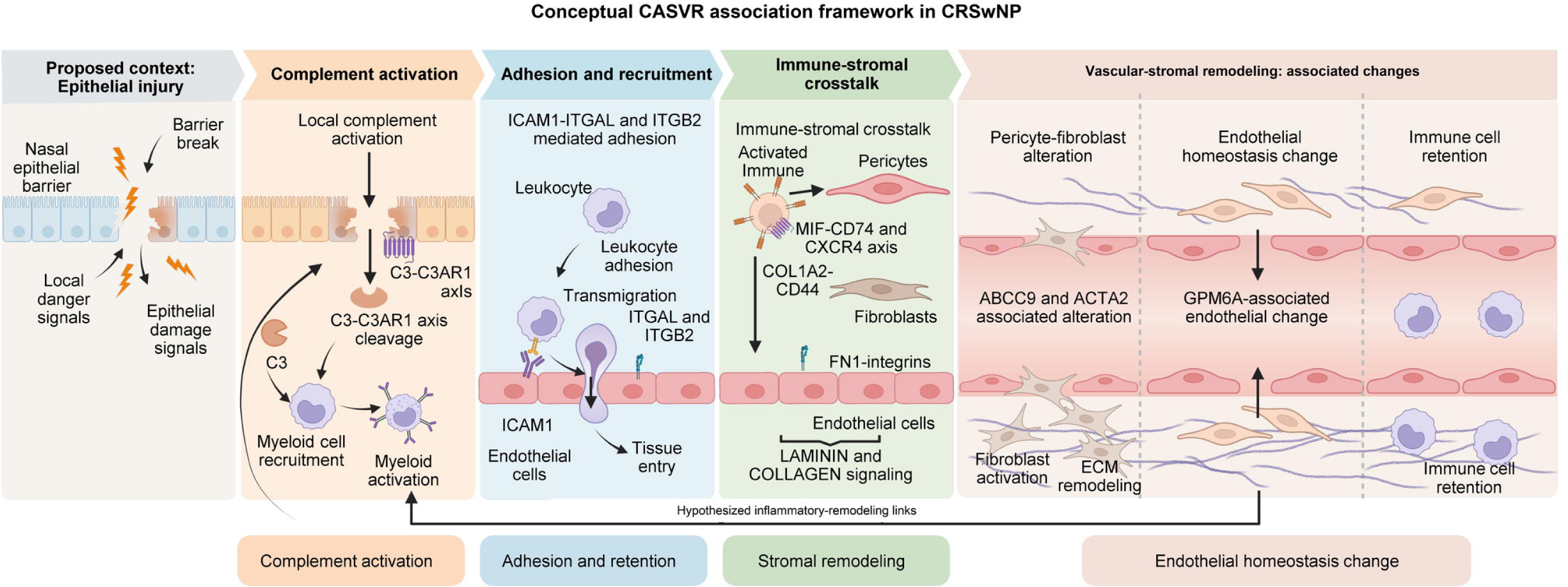
Conceptual CASVR association framework in CRSwNP. The schematic organizes complement-related signals, adhesion-related recruitment, inferred immune-stromal communication, and structural-vascular remodeling into a cross-scale association framework. C3-C3AR1 represents the spatially prioritized complement axis. ICAM1-ITGAL/ITGB2, MIF-CD74/CXCR4, COL1A2-CD44, FN1-integrin, laminin, and collagen represent candidate relationships identified across enrichment, communication, and spatial analyses. Abbreviations: CASVR, complement-adhesion-stromal-vascular remodeling; CRSwNP, chronic rhinosinusitis with nasal polyps; ECM, extracellular matrix.

### Pharmacological mapping supports structure-guided follow-up of C3AR1

To place the spatially prioritized C3AR1 axis and the structural-vascular hub ABCC9 in pharmacological context, we assembled a gene-compound network. The selected associations spanned structural, functional, and perturbational evidence. Anisomycin appeared in both lists through distinct cell-line expression signatures (Figure 8a and Supplementary Data 1 and 2). Cognate-ligand redocking reproduced the deposited SB290157 pose in human C3aR, whereas P1075 in rat SUR2B exceeded the 2.00 Å pose-reproduction criterion (Figure 8b,c and Supplementary Data 3). Only the C3AR1 configuration therefore supported structure-guided follow-up.

**Figure 8.**
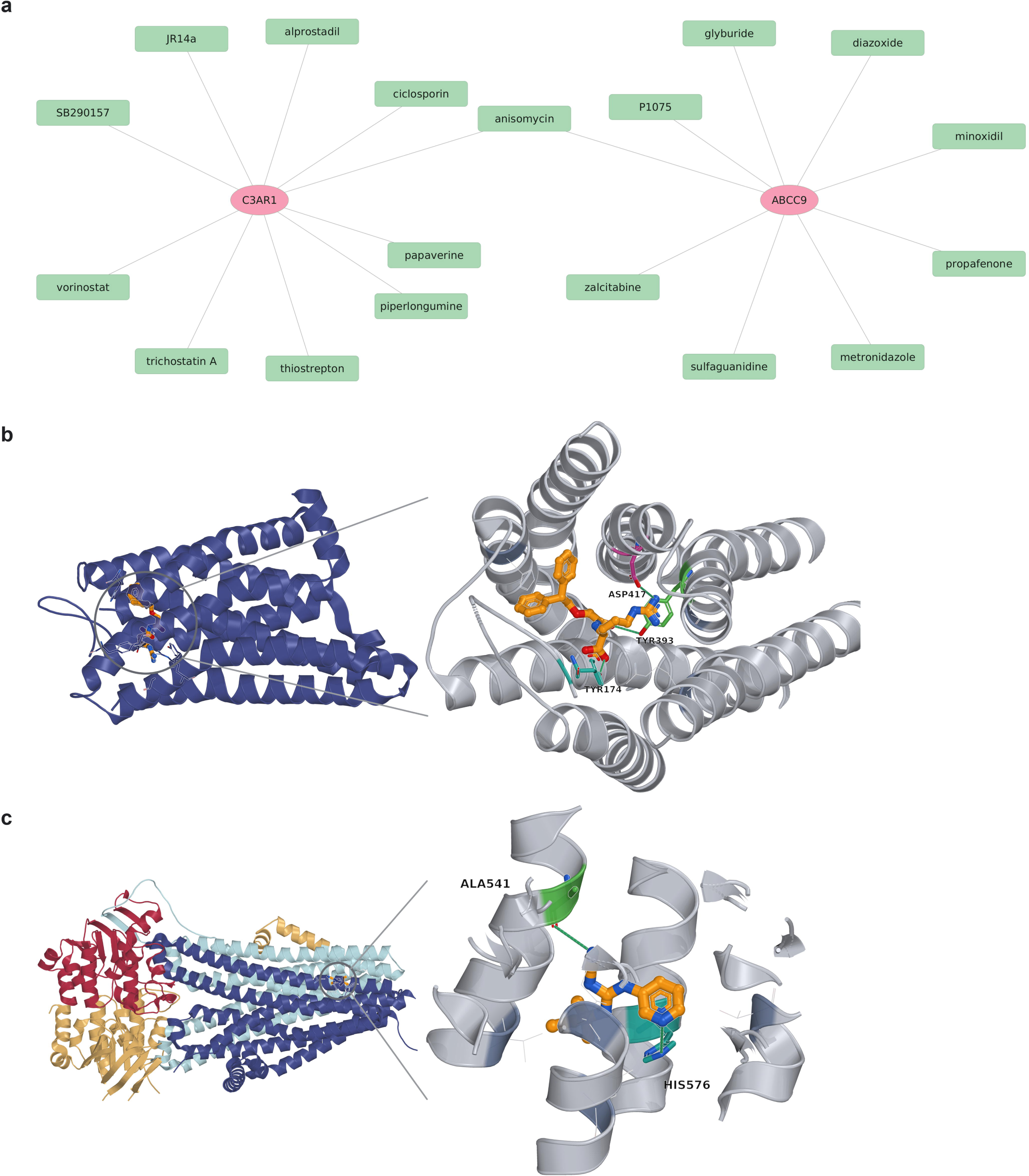
Selected gene-compound associations and cognate-ligand redocking controls for C3AR1 and ABCC9. (a) Selected C3AR1- and ABCC9-associated compounds from DSigDB and curated structural or functional sources. Evidence assignments are listed in Supplementary Data 2. The central anisomycin node represents separate HL60 C3AR1_UP and PC3 ABCC9_UP signatures. (b) Global and enlarged pocket views of SB290157 redocked to human C3aR (PDB 9KVP; RMSD, 1.22 Å). (c) Corresponding views of P1075 redocked to rat SUR2B (PDB 7VLS; RMSD, 2.23 Å), exceeding the 2.00 Å pose-reproduction criterion. Dashed green lines indicate geometric polar contacts separated by 2.3-3.6 Å. Protein backbones are shown as cartoons, ligands as orange sticks, and labelled contact residues in colour.

## Discussion

Across bulk, single-cell, and spatial data, complement-related signals converged with pericyte, fibroblast, and endothelial compartments, while C3-C3AR1 provided the strongest spatially prioritized axis.

CASVR complements established T2 biology. Previous CRSwNP studies emphasized epithelial inflammatory memory, epithelial-immune interactions, and T2-associated remodeling [2,7,8,14,15]. Our cross-scale analysis adds perivascular and stromal compartments to this account. The localization of ABCC9 and ACTA2 to pericytes and fibroblasts, and GPM6A to endothelial cells, links the bulk network to tissue remodeling at the cellular level.

Evidence was stronger for the complement component than for adhesion. Prior studies implicate C3-C3aR signaling in nasal polyps [16,17], and the present spatial analysis prioritized C3-C3AR1 after multiple-testing correction. Prior studies link ICAM1-integrin biology to leukocyte adhesion [18]. Adhesion-related signals appeared in bulk and CellChat analyses, but ICAM1-ITGAL/ITGB2 did not meet the spatial FDR threshold. C3-C3AR1 is therefore the stronger target for follow-up; the adhesion axis remains unresolved. Recent machine-learning studies have reported immune, neutrophil, and metabolic CRSwNP signatures [19–21]. Here, cross-scale network convergence defined the candidate set.

The pharmacological extension further separated the two anchors. The C3aR control reproduced its reference pose, whereas the selected SUR2B configuration did not. C3AR1 therefore provides the stronger starting point for structure-guided follow-up, while ABCC9 remains informative for the pericyte-associated transcriptomic program.

Fibroblasts regulate matrix homeostasis and immune-cell positioning [22–26], whereas pericytes contribute to vascular tone and perivascular homeostasis [27]. CellChat connected these compartments through collagen, laminin, FN1, APP-CD74, COL1A2-CD44, and MIF-CD74/CXCR4 patterns [28–30]. The displayed pairs identify axes for direct testing of how immune recruitment may coincide with stromal and endothelial change.

The three-gene score separated the study-defined groups across the reported cohorts, while same-site and paired analyses preserved the hub-gene directions. We use it as a cross-cohort network readout rather than a calibrated clinical classifier.

The retrospective datasets differed in platform, population, anatomy, and cell composition, and the pooled spatial permutation did not retain patient or segment structure. Matched-anatomy cohorts, patient-aware spatial models, and perturbation of C3-C3AR1 are the clearest next tests.

CASVR integrates these cross-scale associations into a testable model of complement-linked stromal and vascular remodeling in CRSwNP.

## Conclusion

Integration of multi-cohort transcriptomics, machine-learning-based feature selection, single-cell mapping, and pooled AOI-level spatial analysis proposes CASVR as a ideal framework in CRSwNP.

## Methods

### Study design and data sources

Bulk discovery and development used 70 of 103 GSE136825 samples (42 CRSwNP nasal-polyp and 28 control inferior-turbinate tissues; working labels CRSwNP_NP and Control_UT); 33 additional CRSwNP inferior-turbinate samples were used for anatomical-site sensitivity analyses. Control_UT is the source working label for the control inferior-turbinate group denoted Control_IT in Supplementary Information; likewise, source label CRSwNP_UT is denoted CRSwNP_IT in Supplementary Information. Three external bulk-transcriptomic datasets (GSE23552, GSE194282, GSE179265) were used to assess group discrimination across cohorts. Anatomical sites and cellular composition were not uniformly matched or modeled across these cohorts. Single-cell analysis drew from GSE276503, retaining 173,588 cells after decontX, scDblFinder, and standard QC. Exploratory spatial analysis used image-aware GeoMx DSP data. The source spatial table contained 545 segment AOIs across 182 ROIs. After excluding 9 AOIs from 3 Exclude-labeled ROIs, 536 AOIs across 179 ROIs were retained. These comprised 356 CRSwNP AOIs (185 labeled CRSwNP and 171 labeled CRSwNP_UNC), 132 CRSsNP AOIs, 21 Control AOIs, and 27 ImmuneTissue AOIs. All 536 retained AOIs were used for spatial mapping and ligand–receptor proximity analyses; ImmuneTissue AOIs were reported as a separate group. The retained AOIs comprised three predefined GeoMx segment classes: epithelial (EPI; n = 179), immune (IMM; n = 178), and macrophage-enriched (MAC; n = 179).

The datasets used for integrated transcriptomic analysis and evaluation are summarized in Supplementary Table S7.

### Bulk transcriptome analysis

GSE136825 counts were mapped to GENCODE v36 gene symbols, normalized, and log2-transformed where needed. Differential expression was estimated with limma empirical-Bayes models, using |log2 fold change| > 1 and a Benjamini-Hochberg-adjusted P < 0.05.

For the anatomical-site sensitivity analyses, raw counts from all 103 GSE136825 samples were remapped after removal of Ensembl version suffixes, duplicate gene symbols were summed, and each gene-level contrast used filterByExpr, TMM normalization, limma-voom, and robust empirical Bayes moderation. Analysis A compared 33 CRSwNP inferior-turbinate samples with 28 control inferior-turbinate samples. Analysis B used 30 NP-IT pairs that could be matched unambiguously from public sample titles and fit a model with pair and tissue terms; the public metadata did not provide a complete patient-ID key. The five Supplementary Table S8 scores were projected using gene and module scaling parameters frozen in the original 70-sample comparison. Hub-gene FDR values were genome-wide within each contrast, whereas score FDR values were adjusted across five scores (Supplementary Information). Primary-reference estimates in Supplementary Information were re-estimated with this unified raw-count pipeline and may therefore differ slightly from the discovery-analysis values in Supplementary Table S5.

### Functional enrichment, WGCNA, and PPI analysis

GO and KEGG enrichment analyses were performed using clusterProfiler [13] and org.Hs.eg.db. For GSEA, log2FC from the GSE136825 whole-genome differential expression table was used as the sorting variable, with the Hallmark, Reactome, and KEGG_MEDICUS gene sets from MSigDB v2026.1 Hs. Module scores for selected pathways were calculated using Seurat::AddModuleScore on single-cell data.

WGCNA was applied to GSE136825, retaining genes with mean expression > 0.5. A soft threshold of β = 2 was selected, with module identification parameters set to minModuleSize = 30, deepSplit = 2, and pamRespectsDendro = FALSE. Five group-associated modules—red, magenta, purple, green, and royal blue—were selected, and 396 WGCNA candidate genes were filtered using |MM| ≥ 0.6 and |GS| ≥ 0.2. The intersection of these genes with 562 differentially expressed genes yielded 208 candidate contrast-associated network genes.

The 208 candidate genes were imported into STRING [31] to construct a PPI network. In Cytoscape, the top 15 genes were extracted using five cytoHubba algorithms (MCC, MNC, EPC, Stress, and Betweenness). Genes identified by all five algorithms were defined as the final hub genes.

### Machine-learning feature selection and scoring model

Using the eight hub genes (NTRK3, GPM6A, SLITRK5, SPARCL1, ACTA2, RERG, ABCC9, and PDPN) as candidate features, we applied LASSO logistic regression and random forest in GSE136825. LASSO retained ABCC9, ACTA2, and GPM6A, while random forest identified ABCC9, ACTA2, GPM6A, and SLITRK5 using an average importance threshold of 0.125. The intersection retained ABCC9, ACTA2, and GPM6A. These genes were refitted by multivariable logistic regression in all 70 discovery samples to derive the displayed linear predictor. Because candidate generation, feature selection, and refitting used the same cohort without nested resampling, the development AUC is apparent and the fitted probability was not externally calibrated.

The development-set linear predictor is shown with coefficients rounded to three decimal places; analyses used the unrounded coefficients:

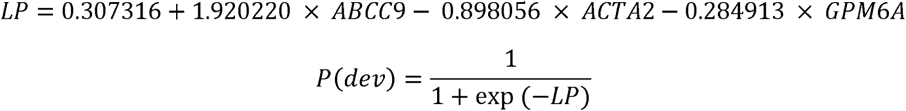

### Single-cell RNA-sequencing analysis

We analyzed the raw GSE276503 10x data using Seurat. We created an object with min.cells = 3 and min.features = 500, used decontX to remove environmental RNA contamination, and retained cells with a contamination score < 0.2. Further quality control criteria included percent_mito < 25, percent_ribo > 3, percent_hb < 5, and nFeature_RNA < 5000. Doublets were removed using scDblFinder.

The post-quality control data were standardized using LogNormalize, and 2,000 highly variable genes were selected. To evaluate the effectiveness of batch correction, we used 26,400 cells selected via balanced sampling to plot UMAP and t-SNE plots obtained via PCA before correction, as well as UMAP and t-SNE plots obtained via Harmony embedding after correction (Supplementary Fig. S2; Supplementary Information). Batch effects were corrected using Harmony based on original identifiers. Clustering and dimensionality reduction were performed using the top 30 dimensions from Harmony, with the FindClusters resolution set to 0.3. We performed manual annotation based on cluster markers and classical lineage markers. We also used UMAP, dot plots, and violin plots to visualize the expression of key genes and pathway module scores.

### Intercellular communication analysis

Intercellular communication was analyzed using CellChat [28] on the integrated single-cell objects via pooled analysis. Stratified random sampling was performed by cell type, retaining up to 1,000 cells per cell type. CellChat objects were created using the RNA data matrix and cell type annotations, with CellChatDB.human as the reference database. The analysis process included subsetData, identifyOverExpressedGenes, identifyOverExpressedInteractions, computeCommunProb, filterCommunication (min.cells=10), computeCommunProbPathway, and aggregateNet. The communication network and key ligand-receptor pairs were visualized using circle plots, signaling role heatmaps, scatter plots, and bubble plots.

### Image-aware spatial transcriptomics analysis

The spatial analysis distinguished predefined GeoMx segment classes—epithelial (EPI), immune (IMM), and macrophage-enriched (MAC)—from four expression-derived functional domains: D2_epithelial, D2_epithelial_c4, D3_stromal_remodeling, and D4_mixed_complement. Spatial expression values were mapped to AOI coordinates for visualization of hub genes and candidate ligand–receptor pairs. Eleven candidate axes were prioritized from bulk enrichment, CellChat-inferred interactions, hub-gene relevance, and prior biological knowledge. All 536 retained AOI records from the three TMA sections and all annotated groups were pooled for this exploratory analysis. Ligand-high and receptor-high AOIs were defined globally as values at or above the upper quartile (top 25%); each high-expression set contained 134 AOIs. Receptor scores were taken from the spatial mapping table, including composite scores for multigene receptor definitions. Euclidean distances were calculated from each ligand-high AOI to its nearest receptor-high AOI, and the mean nearest-neighbor distance was used as the observed statistic. Holding ligand-high labels and coordinates fixed, receptor-high labels were randomly reassigned among AOIs 1,000 times. The one-sided empirical P value was calculated as (number of permuted mean distances less than or equal to the observed distance + 1)/(1,000 + 1), and the proximity z-score as (permutation mean − observed mean)/permutation standard deviation; positive values indicate closer-than-random proximity. Benjamini–Hochberg correction was applied across the 11 candidate axes. FDR < 0.05 was considered significant. The analysis did not constrain distances or permutations by TMA, segment, disease group, ROI, or patient.

### Scoring methods for CASVR and the T2 signature

Supplementary Fig. S3a used per-sample mean single-cell scores rather than count-summed pseudobulks. For CASVR, log-normalized expression was z-standardized gene-wise across all retained cells, signed gene values were averaged within each module, cell-level module scores were averaged within each sample, and the three module means were separately z-standardized across all 33 samples and summed with equal weights. For T2, log-normalized expression of the 14 detected genes was averaged within each sample, z-standardized gene-wise across samples, and then averaged across genes; EPX was not detected in the single-cell object. The IFN-inflammatory score used the same cell-first, sample-mean procedure for IFITM2, HLA-G, SOCS3, OASL, RNF213, HLA-DRB1, IFI27, CCL2, CCL7, MARCO, CCR7, IL18RAP, MMP9, and CXCL13.

For the bulk analysis in Supplementary Fig. S3b, Ensembl version suffixes were removed before GENCODE v36 symbol mapping, duplicate symbols were summed at the count level, and the 42 CRSwNP nasal-polyp and 28 control inferior-turbinate samples were TMM-normalized and converted to log2 counts per million. All 35 CASVR genes were retained. Gene-wise z scores were calculated across the 70 samples; the complement, adhesion, and signed stromal-vascular remodeling modules were calculated as specified in Supplementary Table S8, separately standardized across the same samples, and summed with equal weights. The 42 CRSwNP nasal-polyp samples were then divided at their within-case median into CASVR-high and CASVR-low groups (n = 21 each). High-versus-low statistics were estimated using limma-voom, and moderated t statistics were used for fgsea against Hallmark and Reactome v2026.1 gene sets (minSize = 10; maxSize = 300; seed = 20260818). BH correction was applied across all 1,381 tested pathways; Supplementary Fig. S3b shows the six lowest-FDR pathways in each NES direction.

The gene composition and abbreviations of the CASVR modules and T2 signature are provided in Supplementary Table S8.

Formula:

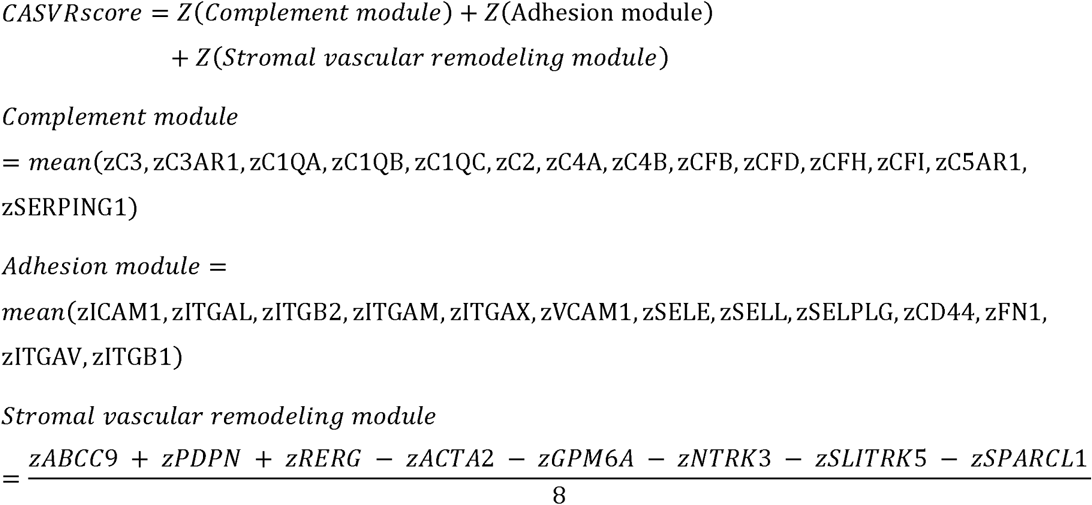

### Gene-compound association analysis and cognate-ligand redocking

C3AR1 and ABCC9 were queried separately in the Drug Signatures Database (DSigDB) [32] on 30 August 2026. Each record was retained with its source annotation, and compound names were deduplicated by exact match. Associations were classified as structural pharmacology, functional or target association, or perturbational expression evidence. A selected network was visualized in Cytoscape 3.10.3 [33]; complete records and displayed edges are provided in Supplementary Data 1 and 2.

Known-ligand redocking assessed pose reproduction independently of the DSigDB associations. Protein chains C and A were extracted from PDB entries 9KVP and 7VLS with Gemmi 0.7.5, and prepared PDBQT files were generated with Meeko 0.8.0. SB290157 was redocked to human C3aR (PDB 9KVP) [34], and P1075 to rat SUR2B (PDB 7VLS) [35]. AutoDock Vina 1.2.7 [36] used deposited-ligand-centred boxes of 22 × 22 × 22 Å and 20 × 20 × 20 Å, respectively, with exhaustiveness 32 and 20 output modes. The three local runs used seeds 20260830, 20260831, and 20260832; the seed-20260830 calculation was repeated in a Linux environment. Top-ranked poses were compared with the deposited ligand by chemical-graph atom mapping without post-docking realignment; Root-mean-square deviation (RMSD) < 2.00 Å was used as the pose-reproduction criterion. The seed-20260831 poses were visualized in PyMOL 3.1.0. Geometric polar contacts were defined as nonredundant protein-ligand N/O/S pairs separated by 2.3-3.6 Å. Complete settings and results are provided in Supplementary Data 3.

### Statistical analysis

We used the Benjamini-Hochberg method for multiple-comparison correction. Unless otherwise specified, an adjusted P < 0.05 or FDR < 0.05 was considered significant. Pearson correlation analysis was used to assess the correlation between WGCNA modules and phenotypes. For Supplementary Fig. S3a, two-sided Wilcoxon rank-sum tests compared CRSwNP (n = 15) with HC (n = 2) for the six displayed scores: CASVR, complement, adhesion, stromal-vascular remodeling, IFN-inflammatory, and T2; BH correction was applied across these six tests. Cohen’s d was calculated as the difference in group means divided by the pooled within-group standard deviation. Sample-level discrimination of the three-gene score was evaluated using the AUC of the ROC curve. Unless otherwise specified, statistical tests were two-sided; the spatial proximity analysis used a one-sided lower-tail permutation test.

## Ethics statement

No new human participants were recruited, no new human specimens were obtained, and no intervention was performed for this study. The publicly available datasets were accessed for research purposes between 1 March 2026 and 3 June 2026. The authors did not have access to information that could identify individual participants during or after data collection; therefore, additional ethics approval and informed consent were not required for this secondary analysis.

## Code availability

The analysis code used in this study is available from the corresponding authors upon reasonable request.

## Use of generative artificial intelligence

Generative artificial intelligence was used solely for language editing and manuscript preparation assistance. The authors verified the resulting text and take full responsibility for all scientific content, analyses, and conclusions.

## Data Availability

The datasets analysed in this study are publicly available in the Gene Expression Omnibus under accession numbers GSE136825, GSE23552, GSE194282, GSE179265, and GSE276503, and in Zenodo at https://doi.org/10.5281/zenodo.13947018. Data needed to evaluate the conclusions are available from these repositories and the accompanying Supplementary Information. Record-level gene-compound associations, displayed network edges, and redocking settings and results are supplied as Supplementary Data 1-3.

https://www.omicsdi.org/dataset/geo/GSE179265

https://www.omicsdi.org/dataset/geo/GSE194282

https://www.omicsdi.org/dataset/geo/GSE276503

https://www.omicsdi.org/dataset/geo/GSE23552

## Acknowledgements

We thank the GEO and Zenodo repositories and the researchers who shared the datasets.

## Author Contributions

Z.-A.M.: Conceptualization, data curation, formal analysis, investigation, methodology, software, visualization, and writing - original draft. X.G.: Methodology, validation, visualization, and writing - review and editing. J.Y.: Methodology, visualization, and writing - review and editing. Y.S.: Conceptualization, resources, supervision, funding acquisition, and writing - review and editing. J.L.: Conceptualization, project administration, resources, supervision, funding acquisition, and writing - review and editing. All authors reviewed and approved the submitted manuscript.

