## Supplementary Information for "Multi-omics proposes a complement-adhesion-stromal-vascular (CASVR) framework in chronic rhinosinusitis with nasal polyps"

Multi-cohort, single-cell, and spatial transcriptomic integration proposes a complement-adhesion-stromal-vascular (CASVR) framework in chronic rhinosinusitis with nasal polyps

Zi-Ang Ma, Xiang Gao, JinLiang Yu, Yu Song and Junxiu Liu

Contents

Supplementary Figures S1-S3 and Supplementary Tables S1-S8. Tables S5-S8 contain the supporting tabular results cited from the main manuscript.

1. Anatomical-site sensitivity analyses

GSE136825 contained 42 CRSwNP nasal-polyp (CRSwNP_NP), 33 CRSwNP inferior-turbinate (CRSwNP_IT), and 28 control inferior-turbinate (Control_IT) RNA-seq samples. Ensembl version suffixes were removed, genes were mapped with GENCODE v36, and duplicate symbols were summed at the raw-count level. Low-expression filtering used edgeR::filterByExpr, followed by TMM normalization, limma-voom, and robust empirical Bayes moderation. The primary contrast was CRSwNP_NP minus Control_IT. Analysis A compared CRSwNP_IT with Control_IT. For Analysis B, the public metadata lacked a patient-ID field; exact numeric matches between PY/SCP and TT/SCT sample titles identified 30 unambiguous NP-IT pairs. These pairs were analyzed with a limma model containing pair and tissue terms. Three CRSwNP_IT samples without an unambiguous public-title match (SCP 201, SCP 213, and SCT 34) were not included in the paired analysis.

The eight hub genes and the five scores specified in Supplementary Table S8 were examined. For score calculation, all 103 samples were TMM-normalized together. Gene and module means and standard deviations estimated in the original 70-sample development comparison were then frozen, and the 33 CRSwNP_IT samples were projected onto this scale. CASVR was the sum of the standardized complement, adhesion, and signed stromal-vascular remodeling modules; the T2 score used the 15-gene Supplementary Table S8 set. Hub-gene P values were adjusted across all expressed genes tested in each contrast (19,239 in the primary contrast, 19,032 in Analysis A, and 18,737 in Analysis B). Score P values were adjusted across the five scores within each contrast. Analysis A effect sizes are Hedges' g; Analysis B effect sizes are paired Cohen's dz. All tests were two-sided. Directional concordance was descriptive and was not assigned a separate P value. Primary-contrast estimates in this appendix were recomputed with the same raw-count pipeline used for Analyses A and B; they are reference estimates for directional comparison and may differ slightly from the discovery-analysis values in Supplementary Table S5.

In Analysis A, all eight hub genes and all five scores changed in the same direction as in the primary CRSwNP_NP-versus-Control_IT contrast. ABCC9 was higher in CRSwNP_IT (log2FC = 1.72; 95% CI, 0.83 to 2.60; genome-wide FDR = 0.020). ACTA2 and GPM6A were lower (log2FC = -0.55 and -0.15), although neither met the genome-wide FDR threshold. The CASVR score was higher by 1.80 units (95% CI, 1.07 to 2.53; five-score FDR = 3.02 × 10−5), and the T2 score was higher by 0.29 units (95% CI, 0.08 to 0.49; FDR = 0.0087). Thus, the same-site comparison retained the overall direction of the primary findings without requiring every selected gene to meet FDR < 0.05.

In the 30 public-title-matched pairs, all eight hub genes and all five scores again changed in the primary direction. ABCC9 was higher in nasal polyps (paired log2FC = 0.87; genome-wide FDR = 0.0064), whereas ACTA2 and GPM6A were lower (paired log2FC = -1.57 and -1.80; FDR = 0.00040 and 0.00039). The paired CASVR difference was 1.89 units (95% CI, 1.35 to 2.43; five-score FDR = 2.20 × 10−7), and the paired T2 difference was 0.37 units (95% CI, 0.18 to 0.56; FDR = 0.00059). These within-CRSwNP results are consistent with a local polyp-associated component, but they do not exclude cellular-composition or other cohort-specific effects.

Supplementary Figure S1

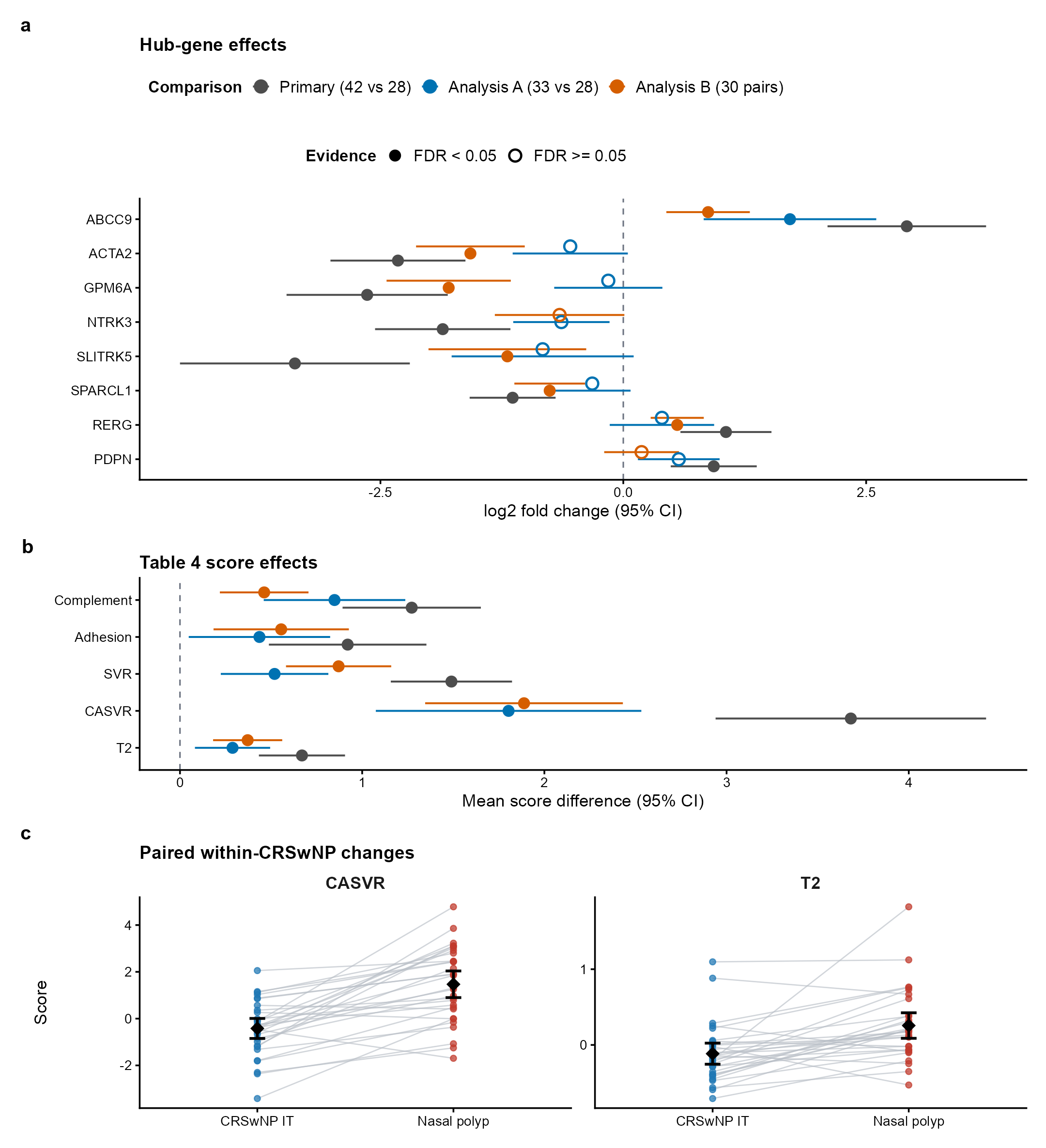

Supplementary Fig. S1. Anatomical-site sensitivity analyses in GSE136825. (a) Hub-gene log2 fold changes and 95% confidence intervals for the primary contrast, Analysis A, and Analysis B. Filled and open points denote genome-wide FDR < 0.05 and FDR ≥ 0.05, respectively. (b) Differences in the five Supplementary Table S8 scores projected onto the frozen 70-sample scale; BH correction covered five scores per contrast. (c) CASVR and T2 scores in 30 public-title-matched pairs; lines join samples and diamonds show means with 95% confidence intervals. IT, inferior turbinate; NP, nasal polyp.

Supplementary Table S1

Supplementary Table S1. Hub-gene estimates in the primary and anatomical-site sensitivity contrasts. Gene-level BH FDR was calculated across all expressed genes shown in the FDR-family column. Primary values are same-pipeline reanalysis estimates for directional comparison and do not replace Supplementary Table S5.

| Gene | Contrast | log2FC (95% CI) | P | BH FDR | FDR family | Direction concordant |
| --- | --- | --- | --- | --- | --- | --- |
| ABCC9 | Primary: NP vs control IT (42 vs 28) | 2.92 (2.10, 3.73) | 6.34E−10 | 1.03E−7 | 19239 | Reference |
| ABCC9 | A: CRSwNP IT vs control IT (33 vs 28) | 1.72 (0.83, 2.60) | 2.69E−4 | 0.020 | 19032 | Yes |
| ABCC9 | B: paired NP vs CRSwNP IT (30 pairs) | 0.87 (0.44, 1.30) | 2.28E−4 | 0.006 | 18737 | Yes |
| ACTA2 | Primary: NP vs control IT (42 vs 28) | -2.32 (-3.01, -1.62) | 4.88E−9 | 4.91E−7 | 19239 | Reference |
| ACTA2 | A: CRSwNP IT vs control IT (33 vs 28) | -0.55 (-1.14, 0.05) | 0.070 | 0.293 | 19032 | Yes |
| ACTA2 | B: paired NP vs CRSwNP IT (30 pairs) | -1.57 (-2.13, -1.01) | 2.45E−6 | 3.99E−4 | 18737 | Yes |
| GPM6A | Primary: NP vs control IT (42 vs 28) | -2.64 (-3.47, -1.81) | 1.84E−8 | 1.33E−6 | 19239 | Reference |
| GPM6A | A: CRSwNP IT vs control IT (33 vs 28) | -0.15 (-0.71, 0.40) | 0.581 | 0.793 | 19032 | Yes |
| GPM6A | B: paired NP vs CRSwNP IT (30 pairs) | -1.80 (-2.44, -1.16) | 2.37E−6 | 3.93E−4 | 18737 | Yes |
| NTRK3 | Primary: NP vs control IT (42 vs 28) | -1.86 (-2.56, -1.16) | 1.12E−6 | 3.14E−5 | 19239 | Reference |
| NTRK3 | A: CRSwNP IT vs control IT (33 vs 28) | -0.64 (-1.13, -0.14) | 0.013 | 0.125 | 19032 | Yes |
| NTRK3 | B: paired NP vs CRSwNP IT (30 pairs) | -0.66 (-1.32, 0.01) | 0.054 | 0.204 | 18737 | Yes |
| SLITRK5 | Primary: NP vs control IT (42 vs 28) | -3.38 (-4.56, -2.20) | 2.49E−7 | 9.76E−6 | 19239 | Reference |
| SLITRK5 | A: CRSwNP IT vs control IT (33 vs 28) | -0.83 (-1.77, 0.11) | 0.081 | 0.315 | 19032 | Yes |
| SLITRK5 | B: paired NP vs CRSwNP IT (30 pairs) | -1.19 (-2.01, -0.38) | 0.005 | 0.047 | 18737 | Yes |
| SPARCL1 | Primary: NP vs control IT (42 vs 28) | -1.14 (-1.58, -0.70) | 2.32E−6 | 5.62E−5 | 19239 | Reference |
| SPARCL1 | A: CRSwNP IT vs control IT (33 vs 28) | -0.32 (-0.71, 0.07) | 0.110 | 0.364 | 19032 | Yes |
| SPARCL1 | B: paired NP vs CRSwNP IT (30 pairs) | -0.76 (-1.12, -0.39) | 1.72E−4 | 0.005 | 18737 | Yes |
| RERG | Primary: NP vs control IT (42 vs 28) | 1.06 (0.59, 1.53) | 2.58E−5 | 3.72E−4 | 19239 | Reference |
| RERG | A: CRSwNP IT vs control IT (33 vs 28) | 0.40 (-0.14, 0.94) | 0.144 | 0.414 | 19032 | Yes |
| RERG | B: paired NP vs CRSwNP IT (30 pairs) | 0.56 (0.28, 0.83) | 2.29E−4 | 0.006 | 18737 | Yes |
| PDPN | Primary: NP vs control IT (42 vs 28) | 0.93 (0.49, 1.37) | 7.72E−5 | 8.74E−4 | 19239 | Reference |
| PDPN | A: CRSwNP IT vs control IT (33 vs 28) | 0.57 (0.15, 0.99) | 0.009 | 0.101 | 19032 | Yes |
| PDPN | B: paired NP vs CRSwNP IT (30 pairs) | 0.19 (-0.20, 0.57) | 0.327 | 0.601 | 18737 | Yes |

Supplementary Table S2

Supplementary Table S2. Supplementary Table S8 score estimates in the primary and anatomical-site sensitivity contrasts. BH FDR was calculated across five scores within each contrast.

| Score | Contrast | Difference (95% CI) | P | BH FDR | Effect size | Direction concordant |
| --- | --- | --- | --- | --- | --- | --- |
| Complement | Primary: NP vs control IT (42 vs 28) | 1.27 (0.89, 1.65) | 4.35E−9 | 7.25E−9 | 1.60 (Hedges g) | Reference |
| Complement | A: CRSwNP IT vs control IT (33 vs 28) | 0.85 (0.46, 1.24) | 4.92E−5 | 1.23E−4 | 1.10 (Hedges g) | Yes |
| Complement | B: paired NP vs CRSwNP IT (30 pairs) | 0.46 (0.22, 0.71) | 4.95E−4 | 6.19E−4 | 0.72 (paired Cohen dz) | Yes |
| Adhesion | Primary: NP vs control IT (42 vs 28) | 0.92 (0.49, 1.35) | 6.47E−5 | 6.47E−5 | 1.01 (Hedges g) | Reference |
| Adhesion | A: CRSwNP IT vs control IT (33 vs 28) | 0.44 (0.05, 0.82) | 0.028 | 0.028 | 0.56 (Hedges g) | Yes |
| Adhesion | B: paired NP vs CRSwNP IT (30 pairs) | 0.56 (0.18, 0.93) | 0.005 | 0.005 | 0.54 (paired Cohen dz) | Yes |
| SVR | Primary: NP vs control IT (42 vs 28) | 1.49 (1.16, 1.82) | 2.97E−13 | 7.43E−13 | 2.16 (Hedges g) | Reference |
| SVR | A: CRSwNP IT vs control IT (33 vs 28) | 0.52 (0.22, 0.81) | 8.24E−4 | 0.001 | 0.89 (Hedges g) | Yes |
| SVR | B: paired NP vs CRSwNP IT (30 pairs) | 0.87 (0.58, 1.16) | 6.74E−7 | 1.68E−6 | 1.11 (paired Cohen dz) | Yes |
| CASVR | Primary: NP vs control IT (42 vs 28) | 3.68 (2.94, 4.42) | 5.00E−15 | 2.50E−14 | 2.34 (Hedges g) | Reference |
| CASVR | A: CRSwNP IT vs control IT (33 vs 28) | 1.80 (1.07, 2.53) | 6.03E−6 | 3.02E−5 | 1.23 (Hedges g) | Yes |
| CASVR | B: paired NP vs CRSwNP IT (30 pairs) | 1.89 (1.35, 2.43) | 4.40E−8 | 2.20E−7 | 1.24 (paired Cohen dz) | Yes |
| T2 | Primary: NP vs control IT (42 vs 28) | 0.67 (0.43, 0.91) | 3.08E−7 | 3.85E−7 | 1.40 (Hedges g) | Reference |
| T2 | A: CRSwNP IT vs control IT (33 vs 28) | 0.29 (0.08, 0.49) | 0.007 | 0.009 | 0.73 (Hedges g) | Yes |
| T2 | B: paired NP vs CRSwNP IT (30 pairs) | 0.37 (0.18, 0.56) | 3.55E−4 | 5.92E−4 | 0.78 (paired Cohen dz) | Yes |

Supplementary Table S3

Supplementary Table S3. Public-title-matched sample pairs used in Analysis B. Pairing was inferred only when the numeric identifiers in PY/SCP or TT/SCT titles matched unambiguously.

| **Pair** | **NP GSM** | **NP title** | **NP BAM** | **IT GSM** | **IT title** | **IT BAM** |
| --- | --- | --- | --- | --- | --- | --- |
| P01 | GSM4058752 | PY 1 | RHP5630.bam | GSM4058762 | SCP 1 | RHP5640.bam |
| P02 | GSM4058753 | PY 10 | RHP5631.bam | GSM4058763 | SCP 10 | RHP5641.bam |
| P03 | GSM4058754 | PY 14 | RHP5632.bam | GSM4058764 | SCP 14 | RHP5642.bam |
| P04 | GSM4058759 | PY 154 | RHP5637.bam | GSM4058768 | SCP 154 | RHP5646.bam |
| P05 | GSM4058760 | PY 155 | RHP5638.bam | GSM4058769 | SCP 155 | RHP5647.bam |
| P06 | GSM4058761 | PY 156 | RHP5639.bam | GSM4058770 | SCP 156 | RHP5648.bam |
| P07 | GSM4058777 | PY 157 | RHP5695.bam | GSM4058771 | SCP 157 | RHP5649.bam |
| P08 | GSM4058757 | PY 182 | RHP5635.bam | GSM4058766 | SCP 182 | RHP5644.bam |
| P09 | GSM4058778 | PY 202 | RHP5696.bam | GSM4058803 | SCP 202 | RHP5722.bam |
| P10 | GSM4058779 | PY 203 | RHP5697.bam | GSM4058804 | SCP 203 | RHP5723.bam |
| P11 | GSM4058780 | PY 204 | RHP5698.bam | GSM4058805 | SCP 204 | RHP5724.bam |
| P12 | GSM4058782 | PY 206 | RHP5700.bam | GSM4058806 | SCP 206 | RHP5725.bam |
| P13 | GSM4058785 | PY 209 | RHP5703.bam | GSM4058807 | SCP 209 | RHP5726.bam |
| P14 | GSM4058786 | PY 210 | RHP5704.bam | GSM4058808 | SCP 210 | RHP5727.bam |
| P15 | GSM4058788 | PY 212 | RHP5706.bam | GSM4058809 | SCP 212 | RHP5729.bam |
| P16 | GSM4058758 | PY 404 | RHP5636.bam | GSM4058767 | SCP 404 | RHP5645.bam |
| P17 | GSM4058756 | PY 75 | RHP5634.bam | GSM4058765 | SCP 75 | RHP5643.bam |
| P18 | GSM4058789 | TT 18 | RHP5707.bam | GSM4058811 | SCT 18 | RHP5731.bam |
| P19 | GSM4058790 | TT 19 | RHP5708.bam | GSM4058812 | SCT 19 | RHP5732.bam |
| P20 | GSM4058791 | TT 20 | RHP5709.bam | GSM4058813 | SCT 20 | RHP5733.bam |
| P21 | GSM4058792 | TT 21 | RHP5710.bam | GSM4058814 | SCT 21 | RHP5734.bam |
| P22 | GSM4058793 | TT 22 | RHP5711.bam | GSM4058815 | SCT 22 | RHP5735.bam |
| P23 | GSM4058794 | TT 23 | RHP5712.bam | GSM4058816 | SCT 23 | RHP5736.bam |
| P24 | GSM4058795 | TT 28 | RHP5713.bam | GSM4058817 | SCT 28 | RHP5737.bam |
| P25 | GSM4058796 | TT 29 | RHP5714.bam | GSM4058818 | SCT 29 | RHP5738.bam |
| P26 | GSM4058797 | TT 30 | RHP5715.bam | GSM4058819 | SCT 30 | RHP5739.bam |
| P27 | GSM4058798 | TT 31 | RHP5716.bam | GSM4058820 | SCT 31 | RHP5740.bam |
| P28 | GSM4058799 | TT 32 | RHP5717.bam | GSM4058821 | SCT 32 | RHP5741.bam |
| P29 | GSM4058800 | TT 33 | RHP5718.bam | GSM4058822 | SCT 33 | RHP5742.bam |
| P30 | GSM4058801 | TT 35 | RHP5720.bam | GSM4058824 | SCT 35 | RHP5744.bam |

2. Harmony batch-correction plots

To evaluate the effectiveness of batch correction for single-cell RNA sequencing data, we generated comparison plots of UMAP and t-SNE visualizations before and after Harmony correction. This analysis was based on 26,400 cells from balanced sampling, 33 samples, 7,200 cells from CRSIT, 5,600 cells from CRSMT, 12,000 cells from CRSNP, and 1,600 cells from HC. Before correction, UMAP and t-SNE dimensionality reduction were performed based on PCA; after correction, UMAP and t-SNE visualizations were re-performed based on the Harmony embedding, with cells colored according to sample origin (orig.ident).

The results show that after Harmony correction, cells from different sample sources are more uniformly mixed in the main low-dimensional spatial structure, with no obvious separation dominated by a single sample, and the spatial structure of the main cell clusters is still preserved. These results demonstrate that Harmony can effectively reduce batch effects between samples, laying a solid data foundation for subsequent cell clustering, cell annotation, and downstream comparative analysis.

Supplementary Figure S2

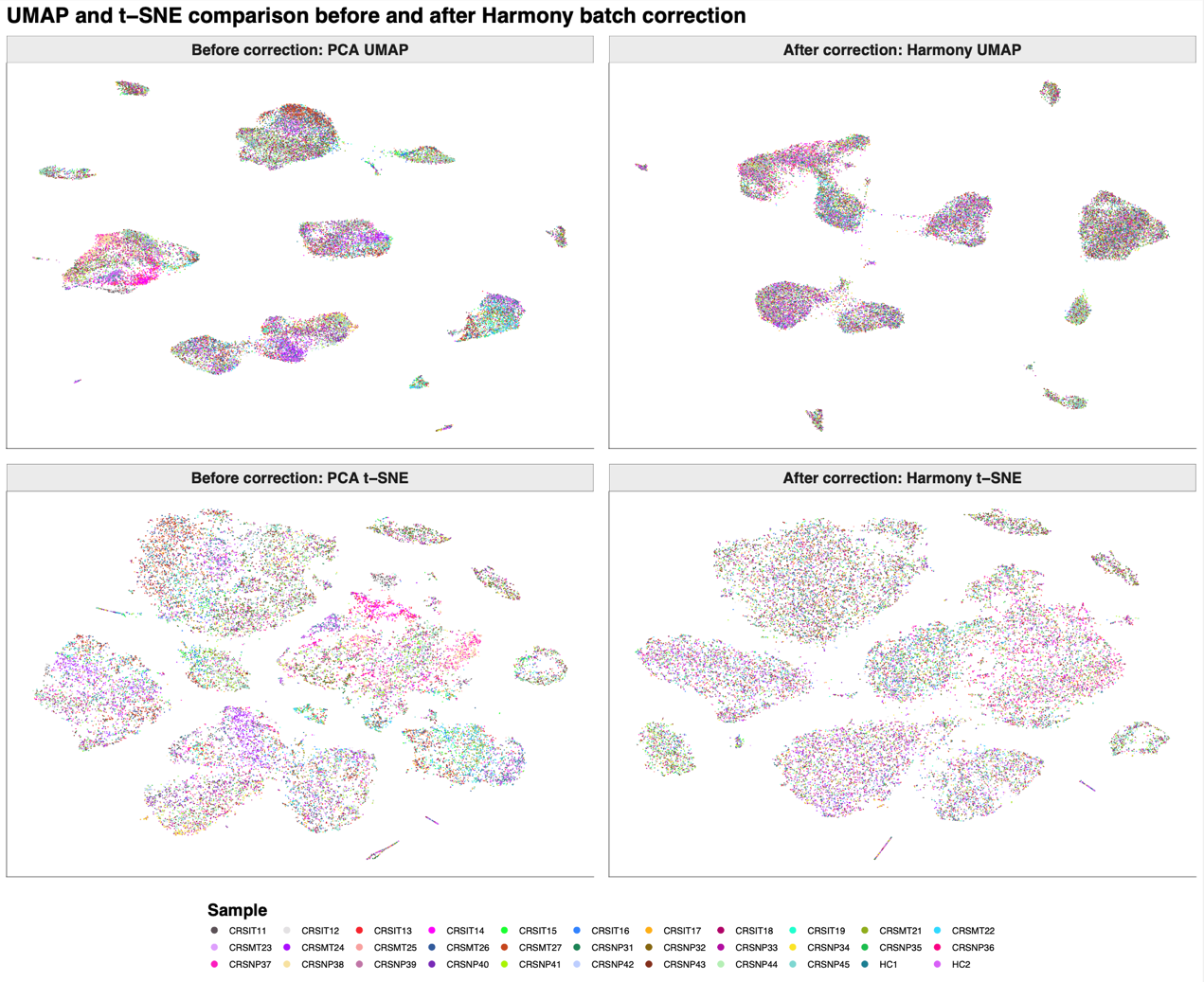

Supplementary Figure S2. Harmony performs batch correction on the single-cell RNA sequencing data before and after, and then compares the UMAP and t-SNE plots. The cells are colored according to their sample origin, showing the distribution of UMAP and t-SNE based on PCA before correction and the distribution based on Harmony after correction.

3. CASVR scores and score-defined pathway enrichment

Supplementary Figure S3

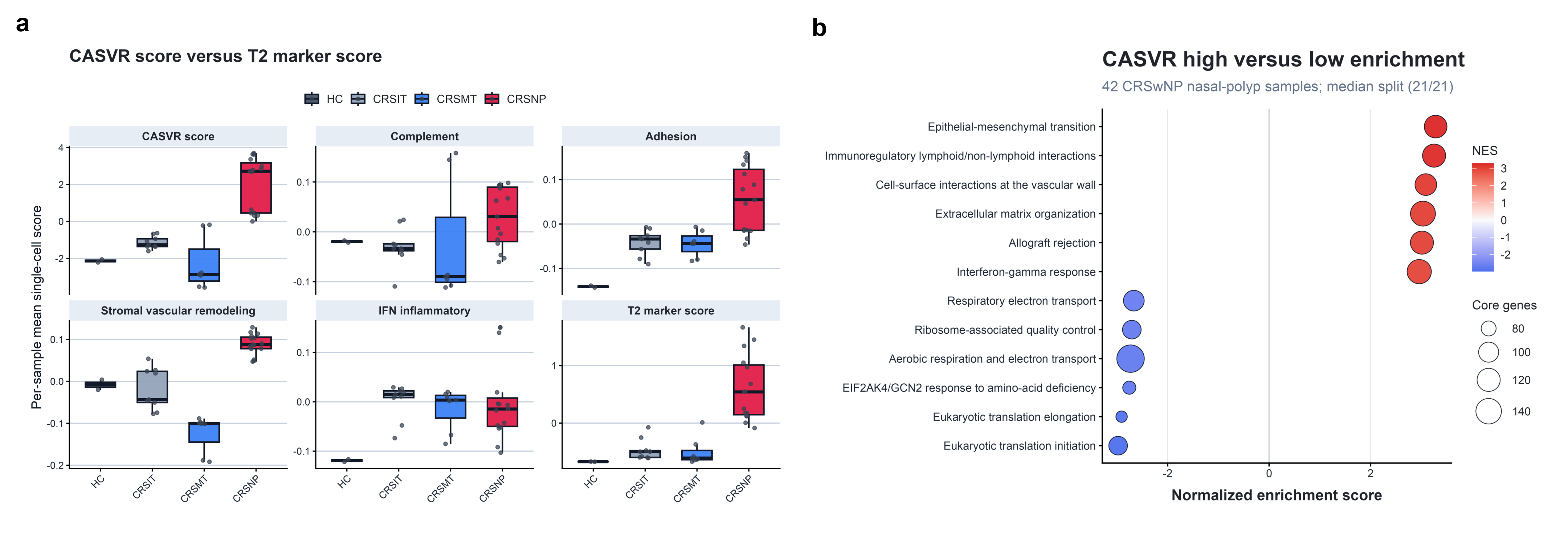

Supplementary Figure S3. CASVR-related scores and score-defined pathway enrichment. (a) Box-and-whisker plots show per-sample mean single-cell CASVR and component scores, together with IFN-inflammatory and T2 scores, across 33 samples (CRSIT, n = 9; CRSMT, n = 7; CRSNP, n = 15; HC, n = 2). For the anatomically unmatched CRSNP-versus-HC comparison, two-sided Wilcoxon P = 0.0147 and BH-adjusted FDR = 0.0176 for both CASVR and T2; Cohen's d was 2.97 and 2.31, respectively. BH correction covered the six displayed scores. (b) Hallmark and Reactome GSEA comparing 21 CASVR-high and 21 CASVR-low GSE136825 CRSwNP nasal-polyp samples, divided at the within-case median of the Supplementary Table S8 CASVR score. Positive NES indicates enrichment in CASVR-high samples and negative NES indicates enrichment in CASVR-low samples. Points show the six lowest-FDR pathways in each direction from 1,381 tested pathways; point size denotes core genes and color denotes NES. The analysis is exploratory and was performed in the score-development dataset. Abbreviations: CASVR, Complement-Adhesion-Stromal-Vascular Remodeling; T2, Type 2; IFN, Interferon; CRSwNP, Chronic rhinosinusitis with nasal polyps; CRSNP, Chronic rhinosinusitis nasal polyp samples; CRSIT, Chronic rhinosinusitis inferior turbinate samples; CRSMT, Chronic rhinosinusitis middle turbinate samples; HC, Healthy control; NES, Normalized enrichment score; FDR, False discovery rate.

4. Spatial ligand-receptor proximity analysis

The exploratory spatial analysis pooled all 536 retained AOI records across three TMA sections and all annotated groups. Eleven candidate ligand–receptor axes were evaluated. Ligand-high and receptor-high AOIs were defined globally using the upper quartile (top 25%) of ligand expression and the predefined receptor score, yielding 134 ligand-high and 134 receptor-high AOIs for each axis. The observed statistic was the mean Euclidean distance from each ligand-high AOI to its nearest receptor-high AOI. Receptor-high labels were randomly reassigned among all AOIs 1,000 times while ligand labels and coordinates were held fixed. One-sided empirical lower-tail P values were calculated from the permutation distribution, and Benjamini–Hochberg correction was applied across the 11 candidate axes. FDR < 0.05 was considered significant; FDR < 0.10 was considered supportive but non-significant.

C3–C3AR1 showed the strongest closer-than-random proximity (z = 2.60; empirical P = 0.004; FDR = 0.044) and was the only axis supported at FDR < 0.05. ICAM1–ITGAL/ITGB2 ranked second (z = 2.20; empirical P = 0.011; FDR = 0.060) and was treated as a supportive but non-significant trend. No other axis met FDR < 0.10. Because distances and label permutations were not constrained by TMA, segment, disease group, ROI, or patient—and segments sharing ROI coordinates were analyzed as separate AOIs—these pooled results may be influenced by array layout and AOI composition and should be interpreted as exploratory rather than disease-specific or patient-level spatial validation.

Supplementary Table S4

Supplementary Table S4. Upper-quartile (top 25%) nearest-neighbor permutation results for 11 candidate ligand–receptor axes; distances are reported in coordinate units.

| **Pair** | **Ligand-high n** | **Receptor-high n** | **Observed mean** | **Permutation mean** | **z** | **Empirical P** | **BH FDR** |
| --- | --- | --- | --- | --- | --- | --- | --- |
| C3–C3AR1 | 134 | 134 | 1217.23 | 1674.32 | 2.603 | 0.004 | 0.044 |
| ICAM1–ITGAL/ITGB2 | 134 | 134 | 1218.49 | 1648.82 | 2.201 | 0.011 | 0.060 |
| CXCL8–CXCR1/CXCR2 | 134 | 134 | 1284.45 | 1558.81 | 1.464 | 0.066 | 0.242 |
| HLA-G–LILRB2 | 134 | 134 | 1456.40 | 1669.84 | 1.139 | 0.137 | 0.301 |
| SPP1–ITGAV/ITGB1 | 134 | 134 | 1415.59 | 1638.22 | 1.083 | 0.132 | 0.301 |
| ICAM1–ITGAM/ITGB2 | 134 | 134 | 1592.74 | 1703.71 | 0.564 | 0.277 | 0.435 |
| CD47–SIRPA | 134 | 134 | 1600.69 | 1724.37 | 0.639 | 0.261 | 0.435 |
| CD200–CD200R1 | 134 | 134 | 1692.55 | 1678.20 | -0.074 | 0.543 | 0.747 |
| HLA-G–LILRB4 | 134 | 134 | 1906.95 | 1745.65 | -0.834 | 0.818 | 0.910 |
| MIF–CD74/CXCR4 | 134 | 134 | 1891.45 | 1660.60 | -1.411 | 0.908 | 0.910 |
| SPP1–CD44 | 134 | 134 | 1911.23 | 1646.78 | -1.315 | 0.910 | 0.910 |

Supplementary Table S5

Supplementary Table S5. Cross-platform transcriptomic summary of the eight hub genes. logFC refers to CRSwNP_NP relative to Control_UT and may reflect disease, anatomical site, or cellular composition.

| Gene | logFC | Group | Main cellular source | Dominant functional spatial domain |
| --- | --- | --- | --- | --- |
| ABCC9 | 2.901 | Up | Pericytes, Fibroblasts | D4 mixed complement |
| SLITRK5 | -3.312 | Down | Spatial immune-mixed, low scRNA | D4 mixed complement |
| ACTA2 | -2.256 | Down | Pericytes, Fibroblasts | D3 stromal remodeling |
| GPM6A | -2.686 | Down | Endothelial | D3 stromal remodeling |
| PDPN | 1.088 | Up | Epithelial, Fibroblasts | D4 mixed complement |
| SPARCL1 | -1.073 | Down | Endothelial, Pericytes, Fibroblasts | D3 stromal remodeling |
| NTRK3 | -1.908 | Down | Fibroblasts, Pericytes | D3 stromal remodeling |
| RERG | 1.233 | Up | Epithelial, Ionocytes, vascular-related | D2 epithelial |

Supplementary Table S6

Supplementary Table S6. Sample-level discrimination estimates for the ABCC9-ACTA2-GPM6A score. Threshold-dependent metrics use dataset-specific, post hoc Youden cutoffs and do not represent performance at a locked threshold. N denotes tissue samples; participant clustering was not modeled in GSE23552.

| **Dataset** | **Cohort type** | **N** | **Positive, Negative** | **AUC (95% CI)** | **Youden cutoff** | **Sensitivity** | **Specificity** | **Accuracy** |
| --- | --- | --- | --- | --- | --- | --- | --- | --- |
| GSE136825 | Development cohort | 70 | 42, 28 | 0.973 (0.921-1.000) | 0.445 | 0.976 | 0.964 | 0.971 |
| GSE23552 | External evaluation | 34 | 21, 13 | 0.993 (0.975-1.000) | 0.243 | 0.952 | 1.000 | 0.971 |
| GSE194282 | External evaluation | 14 | 7, 7 | 0.959 (0.864-1.000) | 0.988 | 0.857 | 1.000 | 0.929 |
| GSE179265 | External evaluation | 24 | 17, 7 | 0.966 (0.897-1.000) | 0.978 | 0.941 | 1.000 | 0.958 |

Supplementary Table S7

Supplementary Table S7. Summary of datasets used for integrated transcriptomic analysis and evaluation. Sampling sites and cellular composition were not uniformly matched; external AUCs describe group discrimination only.

| **Dataset** | **Data type** | **Role** | **Samples** | **Groups** | **Key output** |
| --- | --- | --- | --- | --- | --- |
| GSE136825 | Bulk RNA-seq | Discovery, development, and sensitivity analyses | 103 total (70 discovery/development; 33 sensitivity) | CRSwNP_NP 42; CRSwNP_IT 33; Control_IT 28 | 562 DEGs; WGCNA, PPI; three-gene score; anatomical-site sensitivity analyses |
| GSE23552 | Bulk transcriptome | External evaluation | 34 | CRSwNP 21; Control 13 | AUC 0.993 |
| GSE194282 | Bulk transcriptome | External evaluation | 14 | CRSwNP_NP 7; Control_UT 7 | AUC 0.959 |
| GSE179265 | Bulk transcriptome | External evaluation | 24 | CRSwNP 17; Control 7 | AUC 0.966 |
| GSE276503_RAW | Single-cell RNA-seq | Cellular source, module scores, and CellChat | 173,588 cells | CRSIT 40,807; CRSMT 46,346; CRSNP 77,261; HC 9,174 | 13 annotated cell types; CellChat 2,771 interactions |
| Zenodo image-aware spatial cohort | Spatial transcriptomics + ROI images | Exploratory spatial analysis | 536 retained segment AOIs (545 source; 9 Exclude removed) | CRSwNP 356; CRSsNP 132; Control 21; ImmuneTissue 27 | 3 segment classes; 4 functional domains; spatial ligand-receptor evidence |

Supplementary Table S8

Supplementary Table S8. Gene composition and abbreviations of CASVR modules and the T2 signature

| **Abbreviation** | **Full English name** | **Score module name** | **No. genes** | **Gene symbols** | **Scoring direction** |
| --- | --- | --- | --- | --- | --- |
| C | Complement module | Complement_module | 14 | C3; C3AR1; C1QA; C1QB; C1QC; C2; C4A; C4B; CFB; CFD; CFH; CFI; C5AR1; SERPING1 | Positive (+) |
| A | Adhesion module | Adhesion_module | 13 | ICAM1; ITGAL; ITGB2; ITGAM; ITGAX; VCAM1; SELE; SELL; SELPLG; CD44; FN1; ITGAV; ITGB1 | Positive (+) |
| SVR | Stromal-vascular remodeling module | SVR_module | 8 | Positive genes: ABCC9; PDPN; RERG. Negative genes: ACTA2; GPM6A; NTRK3; SLITRK5; SPARCL1 | Mixed (+ and -) |
| T2 | Type 2 inflammatory signature | T2_signature | 15; 14 scRNA | IL4; IL5; IL13; CLC; POSTN; ALOX15; CCL26; CCL24; IL1RL1; CPA3; MS4A2; CCR3; PRG2; EPX; SIGLEC8 | Positive (+) |
